# Enhancing sleep in professional rugby players: observation and sleep interventions

**DOI:** 10.1101/2025.02.19.25322522

**Authors:** Mathias Goldberg, Arnaud Boutin, Benoit Pairot de Fontenay, Joffrey Cohn, Valentin Michel, Emeric Stauffer, Ursula Debarnot

## Abstract

Sleep is a crucial factor in recovery and must be integrated into athletes’ training plans for optimal performance and well-being. Although professional athletes are advised to sleep for at least 8 hours, many experience shorter sleep durations or poor sleep quality. Sleep interventions have been recently proposed to improve sleep, but their effects remain unclear. This ecological study aimed to evaluate the sleep of a rugby team and to assess the effects of sleep interventions, including sleep hygiene education and relaxation techniques. Thirty-six male professional rugby players were evaluated during two pre- and post-intervention match weeks using objective (actimetric) and subjective (questionnaires) assessments. At baseline, 34 on 36 athletes slept less than 8h per night. Combining both sleep dimensions, 61.1 % of players were considered poor sleepers. After sleep interventions, subjective sleep quality improved (*p* = 0.001, *η*^2^ = 0.22), and athletes went to bed earlier (23:28 ± 00:42 vs. 23:43 ± 00:45 during pre-intervention; *p* = 0.01, *η*^2^ = 0.15). Positive effects of sleep interventions were especially observed among poor sleepers as their objective sleep quantity increased (405.2 min pre-intervention vs. 425.9 min post-intervention *p* = 0.004, *η*^2^ = 0.28). Sleep interventions, composed of theoretical and practical sessions, improved sleep characteristics and might be implemented in athletes’ daily routine. This study offers a simple and accessible method to assess athletes’ sleep, while providing adapted recommendations to optimize or enhance sleep quality and quantity.

**Key points:**

- 61.1% of the rugby players exhibited both poor sleep quality and quantity during the week and weekends.
- Theoretical and practical sleep interventions elicited objective and subjective sleep improvement, especially among poor sleepers.
- Sleep assessment and interventions represent an efficient, and feasible method among team-sport players for improving sleep quality.

## 1. INTRODUCTION

Sleep plays a crucial role in athlete performance and well-being, recognized as a key recovery factor for its physiological and psychological restorative effects (Fullagar et al., 2015; Halson & Juliff, 2017). According to a recent review, 50 to 78 % of athletes experience sleep disturbance, especially after hard training cycles or team-sport matches performed over the weekend (Leduc et al., 2019; Walsh et al., 2021). While general population are advised to sleep more than 7 hours per night (Hirshkowitz et al., 2015), athletes, who should need one hour more sleep to recover properly due to their physical and mental demands, used to sleep regularly less (Lastella et al., 2015; Sargent et al., 2021; Vlahoyiannis et al., 2021). It has been clearly established that poor sleep quality and insufficient sleep quantity have deleterious impacts on decision-making, physical and neurocognitive performance, as well as increasing the risk of illness and injury (Milewski et al., 2014; Simpson et al., 2017). The main causes of sleep disturbances in team-sport athletes are match adrenaline, mental state, blue-light exposure, unfamiliar sleep environments, social jetlag or travel (Walsh et al., 2021). These causes may lead to sleep schedule instability, in particular with a well-known phenomenon of catching up and delaying sleep patterns during weekends (Saidi et al., 2022). Although growing evidence emphasized the crucial need to better consider sleep in athletes, there is still a lack of knowledge using valid and reliable assessment of sleep feasible with team-sport athletes.

For a holistic and relevant examination of sleep, multidimensional assessments are required over a whole week, including both objective and subjective sleep measurements (Buysse, 2014). To objectively assess a large group of team-sport athletes during one or two weeks at the same time, Cunha et al. (2023) recommend to use actigraphy data to evaluate total sleep time (TST) and obtain sleep efficiency (SE), which means the percentage between the time of sleep versus the time in bed. Variations in sleep patterns between weekdays and weekends can also be measured by means of percentages (Coefficient of variation, CV). Alongside objective assessment, sleep can also be evaluated subjectively, providing daily or weekly subjective dimension of sleep (Claudino et al., 2019). For instance, Hooper questionnaire assesses, on a daily basis, the quality, quantity, or latency of sleep based on athletes’ self-perception (Hooper & Mackinnon, 1995) whereas the Athlete Sleep Screening Questionnaire (ASSQ) allows it over the past few days (Bender et al., 2018). ASSQ use is encouraged to identify athletes with sleep disorders using cut-points (Walsh et al., 2021). Prior studies have already considered a single dimension of sleep to categorise their athletes such as Epworth Sleepiness Scale (ESS; Johns, 1991; Pasquier et al., 2023), TST more or less 7 hours (Van Ryswyk et al., 2017), or 50 % of athletes with shorter TST (Caia et al., 2018). To consider the multiple dimensions of sleep, DeSantis et al. (2019) had proposed a 0 to 5 sleep composite score from four objective (sleep quantity, sleep regularity, sleep efficiency, sleep timing) and one subjective (sleep satisfaction) dimensions. However, the subjective dimension seems under-represented in the composite score, with no assessment of sleep latency or quantity. It also appears to be difficult to observe the amplitude of variation over a study with a 0 - 5 score and the TST standards do not correspond to athletes’ needs (i.e. > 6h and < 8h, good sleeper). To date, there is no standard methodology allowing to identify good and poor sleepers among athletes considering the multiple dimensions of sleep to offer sleep tailored interventions.

After assessing objectively and subjectively sleep, sleep interventions can be proposed with the aim of optimizing sleep for good sleepers and enhancing it for poor sleepers. Sleep interventions are a set of approaches used to improve sleep, including sleep education sessions based on sleep hygiene recommendations and relaxation strategies such as cardiac coherence, progressive muscular relaxation, hypnosis, or mental imagery (Bonnar et al., 2018). Caia et al. (2018) tested the effects of two 30-minute sleep education seminars on professional rugby players. The seminars covered topics such as regular sleep patterns, bedtime routines and environment, electronic device use, and caffeine/alcohol consumption before sleep. Rugby players went to bed earlier and woke up later, leading to an increase in TST (+ 20 min; actigraphy). However, these benefits remained temporary, as one month later, players’ sleep behaviour reverted to those adopted before sleep education. Another study evaluated the effect of sleep education program during 6 weeks among football players (Van Ryswyk et al., 2017). After weekly meeting and educational sessions, subjective TST extended and SE increased. Nevertheless, the long-term preservation of these benefits has not been assessed (Van Ryswyk et al., 2017). Further studies had evaluated the effects of specific recommendations such as sleep extension on rugby players or napping on various athletes, and both resulted in TST increase (Swinbourne et al., 2018; Lastella et al., 2021; respectively). More recently, two studies combined sleep education and practical strategies. First, Pasquier et al. (2023) evaluated with actigraphy and questionnaires the effect of napping and three sleep-education sessions, each separated by one month, on swimmers. Somnolent swimmers went to bed earlier resulting in extended time in bed, with a more stable wake-up time schedule. Secondly, Vachon et al. (2023) conducted two educational sessions and administrated three relaxation strategies sessions among young rugby players to improve their sleep quality and facilitate falling asleep. Actigraphy measurements showed a small improvement in sleep onset latency (SOL) and SE, which has had a major impact on poorer sleepers (Vachon et al., 2023). As a result, sleep interventions based on a combination of educational session and relaxation strategies seem effective and applicable with a large group of athletes. The multidimensional evaluation of this type of sleep interventions still needs to be examined, as the long-term preservation of sleep-changes in behaviour.

This study aimed to (i) evaluate the multidimension of sleep among team-sport athletes identifying good and poor sleepers, and (ii) assess the acute and long-term effects of sleep interventions, combining sleep education and relaxation strategies. First, we hypothesized that a majority of professional rugby players experience objective and subjective sleep disturbances, especially in sleeping less than 7 hours. Second, we expected that sleep interventions may improve their TST, especially for poorer sleepers. Third, we explored the remaining effects of sleep interventions over one to three months.

## 2. MATERIALS AND METHODS

### 2.1. Participants

Forty-three male professional rugby players from a national French rugby union were included. Seven rugby players missed one or both weeks due to injury, player’s unavailability, or mishandling of the equipment. Thus, thirty-six rugby players completed the two experimental weeks (27.7 ± 4.6 years, range: 19 – 35; height: 183.8 ± 6.9 cm; body mass: 100.2 ± 13.3 kg; BMI: 29.1 ± 3.1 kg·m^-2^; VO_2Max_: 44.7 ± 6.7 kg^−1^.min^−1^; rugby roles: two fullbacks, four wings, three centres, four fly-halfs, one scrumhalf, five back rows, four locks, four hookers, nine props; Table 1). Participants trained daily during the week and had a match on Saturday evening. They gave written informed consent before taking part in the experiment. This study was conducted in accordance with current national and international laws and regulations governing the use of human subjects (Declaration of Helsinki). The study protocol was approved by the Hospices Civil Lyon’s ethical committee.

**Table 1.** Demographic characteristics of the sample (n = 36).

| | Mean ± SD | Min - Max |
| --- | --- | --- |
| Age (year) | 27.7 ± 4.6 | 19 - 35 |
| Height (cm) | 183.8 ± 6.9 | 171 - 200 |
| Body weight (kg) | 100.2 ± 13.3 | 73 - 126 |
| BMI (kg·m<sup>-2</sup>) | 29.1 ± 3.1 | 24.1 - 35.7 |
| Chronotype - MEQ | 58.2 ± 8.2 | 42 - 74 |
| ESS | 7.0 ± 3.8 | 0 - 16 |
| VO<sub>2Max</sub> (kg<sup>-1</sup>·min<sup>-1</sup>) | 44.7 ± 6.7 | 30.2 - 53.5 |

BMI, body mass index; MEQ, Morningness-Eveningness Questionnaire; ESS, Epworth Sleepiness Scale.

### 2.2. Experimental design

Participants were monitored using both objective and subjective assessments during pre- and post-intervention weeks, separated by a one-month interval (*Figure 1*). Participants were first screened using questionnaires about their general daytime sleepiness (ESS) and chronotype (Morningness-Eveningness Questionnaire; Horne & Östberg, 1976). At the beginning of the PRE-INTERVENTION week, two sleep experts raised participants’ awareness of the importance of sleep for optimal performance and recovery, introduced them to the study, and explained how sleep was recorded (15 min, E.S. - MD/PhD and M.G. - PT/PhD student).

**Figure 1:**
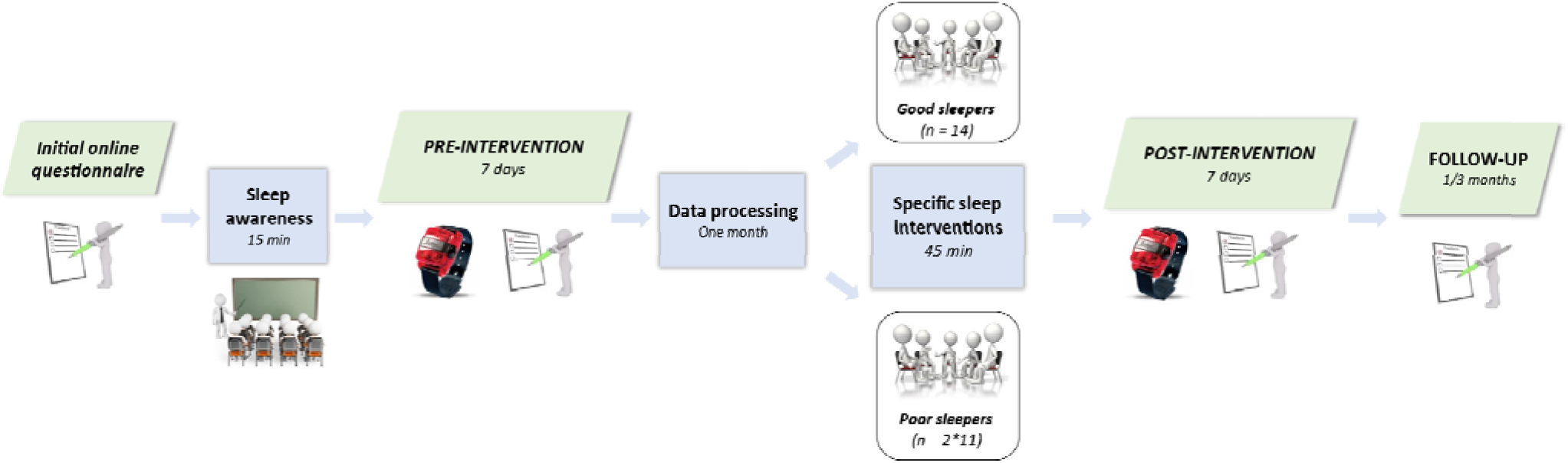
Experimental design. Participants started the study by answering an initial questionnaire. After the sleep awareness session, participants were monitored during one PRE-INTERVENTION week. After one month of data processing, participants received specific sleep interventions in group settings and have been directly reassessed. Long-term effects of sleep intervention were assessed after one and three months.

During the PRE-INTERVENTION week, spanning from Monday to Monday, participants had to maintain their daily habits while continuously wearing an actimeter 24/7 and completing daily online questionnaires sent on their smartphones within the first hour after awaking (i.e., Hooper and questions about screen time exposure and nap). The one-month period between the PRE- and POST-INTERVENTION weeks was dedicated to process the objective and subjective data, whereby participants were categorized as good and poor sleepers using a composite score (detailed below). At the beginning of the POST-INTERVENTION week, the two sleep experts briefly presented the results from the PRE-INTERVENTION to the entire group and insisted on the importance and benefits of sleep, functions of sleep on the human organism, and circadian rhythm. After, rugby players were divided into groups of good sleepers (14 good sleepers) and poor sleepers (two groups of 11 poor sleepers) and had a sleep educational session (30 min) adapted to optimise or enhance sleep in both groups, respectively. Accordingly, sleep experts further recommended to implement sleep hygiene strategies, along with brief theorical explanation, including: 1) sleep extension in the morning, 2) power nap, 3) earlier bedtime, 4) regular wake-sleep cycle, 5) respect individual chronotype, 6) limit screen exposure especially one hour before bedtime, adopt a sleep routine, and 7) avoid evening heavy meals or consume alcohol or coffee. After answering to some questions, sleep experts further invited the participants to experience three relaxation strategies (15 min): cardiac coherence, progressive muscle relaxation, and mental imagery (Jacobson, 1938; Perreault-Pierre, 2012; McCraty & Zayas, 2014; Walker, 2017). After the sleep intervention, participants were given a leaflet synthetizing the sleep recommendations and emphasizing the goal of combining at least two recommendations during the POST-INTERVENTION week. The leaflet also includes a QR code to access audio recordings of the three relaxation strategies, enabling participants to use it if needed during the POST-INTERVENTION week. During this second week, they were monitored using objective and subjective data similar to those from the first week. In addition, two follow-up questionnaires were administrated at one and three months POST-INTERVENTION to assess the long-lasting effects of the intervention, including: 1) the ASSQ, 2) questions about their current wake-up and bedtimes, and 3) the quantification of recommendations application.

### 2.3. Objective and subjective assessments

To objectively evaluate sleep, participants wore an actimeter (GTX3+, ActiGraph, Pensacola, USA) 24/7 for seven consecutive days (during the PRE- and POST-INTERVENTION weeks) on the non-dominant wrist, except during showers and contact trainings. Actilife LLC Pro software v6.13.4 (Actigraph, Pensacola, USA) was used to analyse wake-sleep cycle and sleep data with the Cole-Kripe algorithm. The sleep window, defined as the period between the ‘time in bed’ and ‘time out of bed’ was first determined by the algorithm, followed by visual inspection, and verified in concordance with self-reported bed/wake-up times. In the event of a mismatch between these data, a manual correction of the algorithm data was made in favour of the declared bedtime and wake-up times (DeSantis et al., 2019). All nights of sleep were analysed for the following variables: 1) bedtime (h:min), 2) wake-up time (h:min), 3) total sleep time (min), 4) time in bed (min), 5) sleep efficiency (%), 6) wake after sleep onset (min), and 7) coefficient of variation (CV) of TST. A 60-second epoch sample had been used here.

To subjectively assess sleep, participants completed the ASSQ (Bender et al., 2018) at the beginning of the PRE-INTERVENTION week, at the end of the POST-INTERVENTION week, and after one and three months. From the ASSQ, the sleep difficulty scale (SDS) evaluated sleep parameters with five questions on sleep quantity (0 - 4 scale), sleep quality (0 - 4 scale), sleep latency (0 - 3 scale), nocturnal awakenings (0 - 3 scale), and sleep medicine (0 - 3 scale). A total score equal to or below 4 (out of 17) means good sleep. Participants daily completed the five questions from the Hooper questionnaire (Hooper & Mackinnon, 1995), within the first hour after waking up, assessing fatigue, sleep quality, sleep quantity, muscle soreness, and psychological state; along with an additional query regarding sleep latency question. To specifically consider subjective sleep, we have combined questions on sleep quality (1 – 6 scale), quantity (1 – 7 scale), and latency (1 – 6 scale). This combination, namely the daily subjective sleep, considers a score of 9 or less out of 19 as indicative of good sleep quality. Moreover, daily screen time and nap duration (if applicable) were also reported each day.

### 2.4. Categorisation as good and poor sleepers

With the aim to provide adapted sleep interventions, participants were categorised as good and poor sleepers. As DeSantis et al. (2019), our sleep composite score included objective parameters of sleep efficiency (SE) and regularity (Coefficient of variation of total sleep time [CV_TST_]) and subjective sleep satisfaction including perception of sleep quality and quantity (SDS from the ASSQ and daily subjective sleep). Therefore, the sleep composite score formula used here was as follows:

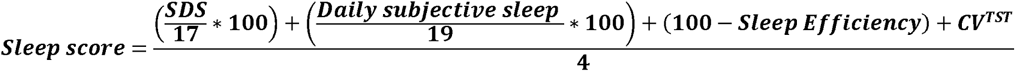

Sleep composite score was calculated at the end of each experimental week, where lower scores indicated good sleep quality. The cut-off between good and poor sleepers was calculated on the basis of the norm of each variable (SDS: 4/17, Daily subjective sleep: 9/19, Sleep Efficiency: 85/100). We decided to consider CV_TST_ = 19.5 %, as cut-off value, based on the prior findings of Saidi et al. (2023) among young elite rugby players. Thus, by applying our sleep composite score formula, an athlete was considered as good sleeper if his score was below 26.35.

### 2.5. Statistical analysis

The primary outcome was the objective TST. Secondary outcomes were each objective sleep variables (bedtime, wake-up time, TIB, SE, WASO and coefficient of variation of TST), subjective sleep variables (SDS, Daily subjective sleep, total score at Hooper questionnaire, daily screen time and nap duration) and the sleep composite score. As residuals followed linearity, homoscedasticity and normality (Shapiro-Wilk test), linear mixed models were implemented to assess the effect of the intervention (PRE-compared to POST-INTERVENTION, to the one- and three-month POST-INTERVENTION follow-up) on primary and secondary outcomes. When the model revealed a main effect, Tukey post-hoc comparisons were performed. Kruskal-Wallis tests were used to evaluate the effect of the day (WEEK DAYS vs. WEEKEND [FRIDAY, SATURDAY]) on bedtime, wake-up time, SE, TST, WASO, Daily subjective sleep and screen time exposure. When the Kruskal-Wallis revealed a main effect, Wilcoxon post-hoc comparisons were performed. The significant threshold was set at 0.05 for all analyses. Effect sizes (*η*^2^) were calculated to interpret the magnitude of the mean difference between the two weeks or days of the week/weekends (small [0.01], medium [0.06] and large [0.14]; Cohen, 1988). Statistical analysis was performed using the R software (version i386 4.1.2).

## 3. RESULTS

### 3.1. PRE-INTERVENTION week

Sleep assessment of the entire group is presented in Table 2. On average, they slept 421.3 min (∼ 7.0 h) per night during the PRE-INTERVENTION week. Results revealed that 16 rugby players slept less than 7 hours per night and 34 on 36 players slept less than 8 hours per night. They went to bed at 23:43 ± 00:45 and woke up at 07:35 ± 00:34. A 15.7 ± 5.4 % of TST variability (CV_TST_) was found between the seven days of the week. On the subjective sleep dimension, nine rugby players had moderate sleep disturbances (score SDS ≤ 8) and 16 had mild sleep disturbances (≤ 5 score SDS ≥ 7). Daily subjective sleep is assessed on average at 10.3 out of 19 (i.e. 1.3 points above the score indicating good sleep). The sleep composite score identified 14 rugby players (mean: 23.2, range: 20.3 – 26.1) as good sleepers and 22 players as poor sleepers (mean: 31.9, range: 27.4 – 39.3). No anthropometric difference were observed between both groups.

**Table 2.** Objective and subjective sleep data throughout the night in PRE-INTERVENTION week for the entire group (PRE; mean and ± SD) and in PRE- and POST-INTERVENTION weeks for good and poor sleepers (POST; mean and ± SD).

|  | ENTIRE GROUPE<br>(n = 36) | GOOD SLEEPERS (n = 14) |  |  | POOR SLEEPERS (n = 22) |  |  |
| --- | --- | --- | --- | --- | --- | --- | --- |
| INTERVENTION | PRE | PRE | POST | *P value* | PRE | POST | *P value* |
| **Subjective variables (scores)** |  |  |  |  |  |  |  |
| ASSQ – SDS | 5.7 ± 2.2 | 4.0 ± 1.7 | 3.8 ± 1.9 | .8 | 6.8 ± 1.8 | 6.5 ± 2.3 | .5 |
| Sleep quality | 3.2 ± 0.8 | 2.6 ± 0.6 | 2.4 ± 0.7 | .4 | 3.7 ± 0.5 | 3.4 ± 0.6 | .08 |
| Sleep quantity | 4.7 ± 0.7 | 4.2 ± 0.6 | 3.7 ± 0.8 | **.007\*\*\*** | 5.1 ± 0.6 | 4.8 ± 0.8 | **.04\*** |
| Sleep latency | 2.3 ± 0.8 | 2.9 ± 0.4 | 2.1 ± 0.8 | .5 | 2.5 ± 0.9 | 2.2 ± 0.9 | .09 |
| Daily subjective sleep | 10.3 ± 1.7 | 8.7 ± 0.9 | 8.3 ± 1.8 | .2 | 11.3 ± 1.3 | 10.5 ± 1.7 | **.003\*\*** |
| Fatigue | 3.8 ± 0.8 | 3.1 ± 0.7 | 3.0 ± 1.1 | .5 | 4.3 ± 0.5 | 4.0 ± 0.8 | .09 |
| Muscle soreness | 3.6 ± 1.1 | 3.0 ± 1.1 | 3.0 ± 1.3 | .9 | 4.0 ± 0.9 | 4.2 ± 0.6 | .2 |
| Psychological state | 3.2 ± 1.0 | 2.6 ± 0.7 | 2.5 ± 1.0 | .7 | 3.6 ± 0.9 | 3.7 ± 0.8 | .8 |
| Hooper total | 21.0 ± 3.8 | 17.4 ± 2.9 | 14.3 ± 3.7 | **<.001\*\*\*** | 23.2 ± 2.5 | 18.8 ± 2.6 | **<.001\*\*** |
| Screen time exposure (min) | 03:53 ± 01:25 | 03:48 ± 01:20 | 04:23 ± 01:23 | **.03\*** | 03:56 ± 01:32 | 03:35 ± 01:26 | .2 |
| **Objectives variables** |  |  |  |  |  |  |  |
| Bedtime (hh:mm) | 23:43 ± 00:45 | 23:33 ± 00:35 | 23:27 ± 00:47 | .5 | 23:48 ± 00:54 | 23:29 ± 00:39 | **.004\*\*** |
| Wake-up time (hh:mm) | 07:35 ± 00:34 | 07:46 ± 00:31 | 07:45 ± 00:31 | .9 | 07:30 ± 00:34 | 07:29 ± 00:37 | .9 |
| Sleep efficiency (%) | 89.4 ± 4.4 | 90.5 ± 3.3 | 88.3 ± 4.5 | **.006\*\*** | 88.7 ± 4.9 | 88.8 ± 4.5 | .8 |
| Total sleep time (min) | 421.3 ± 42.5 | 446.6 ± 40.6 | 441.0 ± 29.9 | .5 | 405.2 ± 36.0 | 425.9 ± 39.1 | **.004\*\*** |
| CV<sub>TST</sub> (%) | 15.7 ± 5.4 | 13.8 ± 5.7 | 11.7 ± 7.4 | .4 | 16.9 ± 4.9 | 14.0 ± 6.4 | .1 |
| Time in bed (min) | 472.5 ± 39.4 | 492.5 ± 31.3 | 499.8 ± 35.9 | .2 | 459.7 ± 39.2 | 480.1 ± 42.0 | **.02\*** |
| Wake after sleep onset (min) | 49.7 ± 20.4 | 45.8 ± 14.2 | 57.8 ± 24.5 | **.01\*\*** | 52.1 ± 23.5 | 53.3 ± 22.7 | .7 |
| Sleep composite score | 28.5 ± 5.3 | 20.3 ± 1.3 | 19.6 ± 4.3 | .5 | 26.9 ± 2.8 | 24.9 ± 4.9 | **.02\*** |

5 CV<sub>TST</sub>, Coefficient of variation of total sleep time. The effects between PRE and POST-INTERVENTION weeks 6 among good and poor sleepers are indicated by “\*” ( $p \leq 0.05$ ), “\*\*” ( $p \leq 0.01$ ) and “\*\*\*” ( $p \leq 0.001$ ). For score 7 variables, a low score means good sleep quality. ASSQ – SDS, Athlete sleep screening questionnaire – Sleep 8 disorder scale.

### 3.2. POST-INTERVENTION

Reported in daily diary, all rugby players implemented at least two recommendations during POST-INTERVENTION week. The effects of sleep interventions on good and poor sleepers are presented in Table 2. Among poor sleepers, a significant positive effect of sleep interventions was found on TST (405.2 min PRE vs. 425.9 min POST; *p* = 0.004, *η*^2^ = 0.28), TIB (*p* = 0.02, *η*^2^ = 0.20), bedtime (*p* = 0.004, *η*^2^ = 0.28), daily subjective sleep (*p* = 0.003, *η*^2^ = 0.29), and Hooper total score (*p* < 0.001, *η*^2^ = 0.82). Overall, the sleep composite score decreases significantly among poor sleepers (*p* = 0.02, *η*^2^ = 0.22) and the entire group (*p* = 0.05, *η*^2^ = 0.10), meaning a multidimensional sleep improvement. Among good sleepers, no significant difference in TST was observed for good sleepers relative to PRE-INTERVENTION. A significant sleep interventions effect was observed on the Hooper total score (*p* < 0.001, *η*^2^ = 0.71), specifically for subjective sleep quantity (*p* = 0.007, *η*^2^ = 0.36). However, SE decreased (*p* = 0.006, *η*^2^ = 0.37) due to an increase in WASO (*p* = 0.01, *η*^2^ = 0.31). No other effect was found between the PRE- and POST-INTERVENTION weeks among good and poor sleepers. Detailed results among the entire group are presented in the supplementary file (Table 1).

### 3.3. Week days and weekend analysis

During the PRE-INTERVENTION week, good sleepers woke up later during the weekend rather than during the week days, inducing greater TST (*p* ≤ 0.001, *η*^2^ = 0.11 and *p* = 0.02, *η*^2^ = 0.04 respectively). No difference was found between week and weekend nights among good sleepers during the POST-INTERVENTION week (Table 2 in supplementary file). During PRE-INTERVENTION week, poor sleepers went to bed and woke up later during weekend rather than weekdays (*p* ≤ 0.001, *η*^2^ = 0.07 and *p* ≤ 0.001, *η*^2^ = 0.15 respectively) inducing no increase in TST (*p* = 0.4) (Table 2 in supplementary file). They conserved the same shift observed in PRE-INTERVENTION week going to bed and waking up later during the weekend rather than weekdays (*p* = 0.007, *η*^2^ = 0.07; *p* = 0.003, *η*^2^ = 0.08 respectively) while TST remained unchanged (*p* = 0.8) during POST-INTERVENTION week (*Figure 2*).

**Figure 2:**
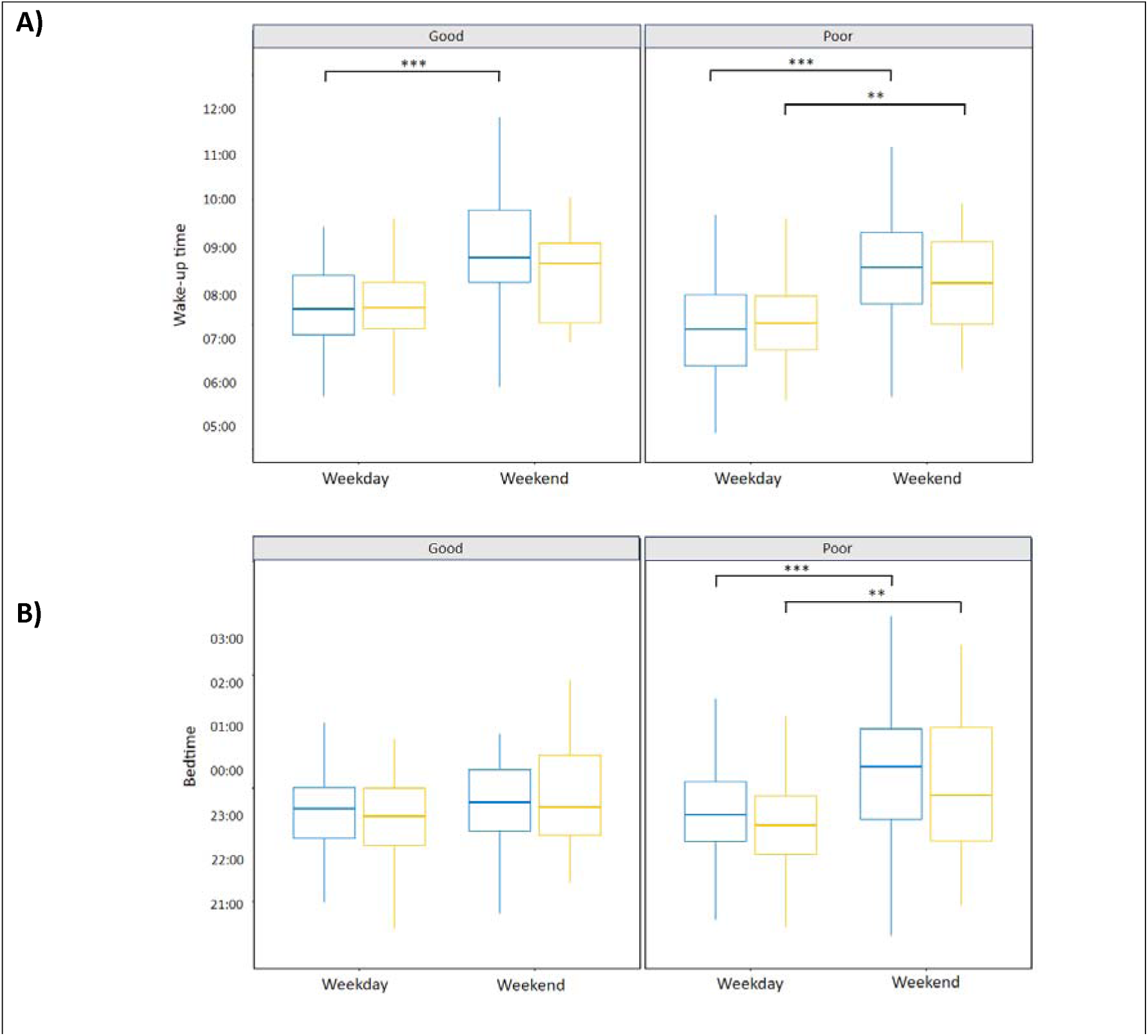
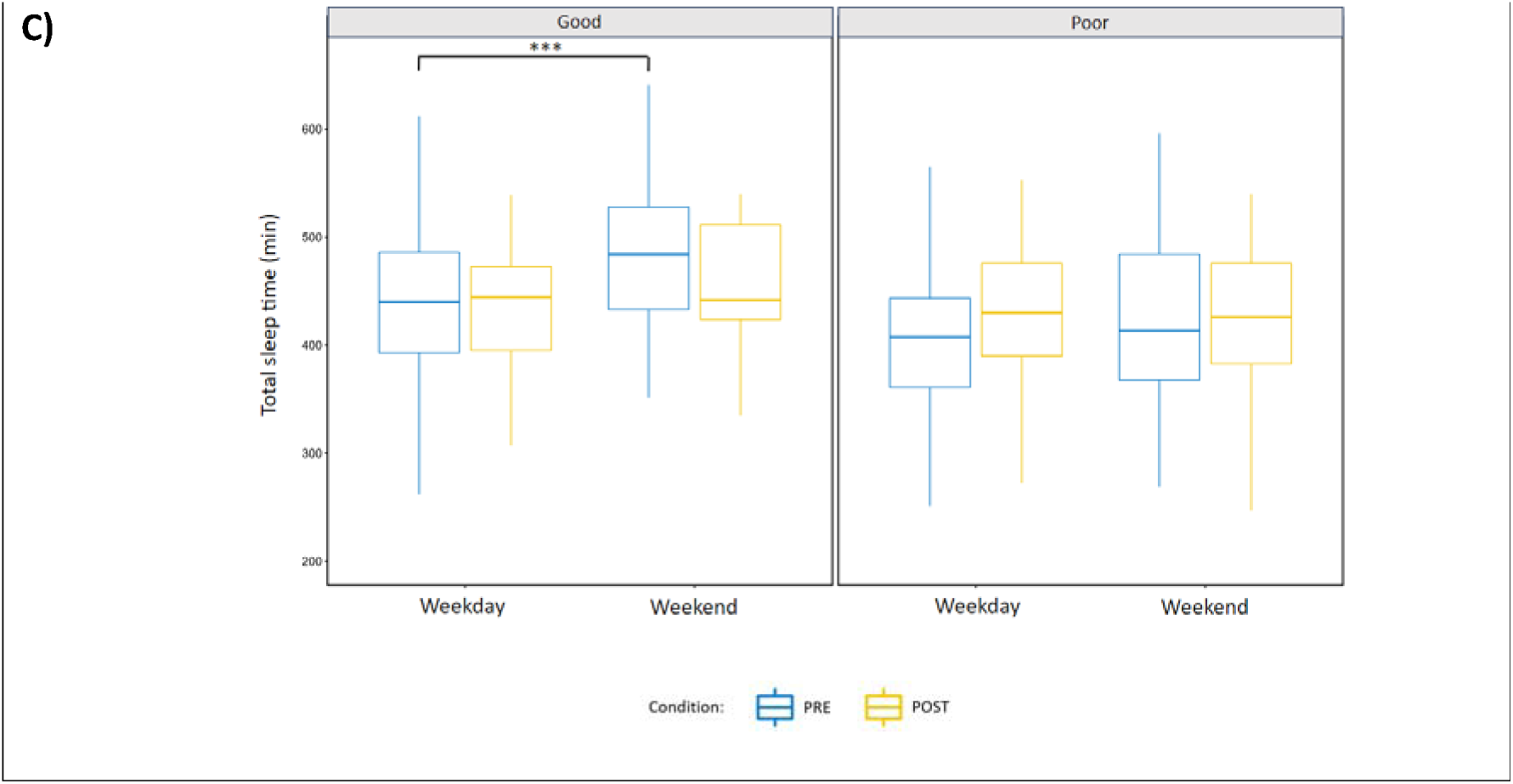
Differences between week and weekend nights (Friday / Saturday) among good and poor sleepers on wake-up time (A), bedtime (B), total sleep time (C) during PRE- (blue) and POST-INTERVENTION (yellow) weeks. (A) Good and poor sleepers woke up later during the weekend compared to weekdays during PRE- and POST-INTERVENTION only for poor sleepers. (B) Poor sleepers went to bed later during the weekend compared to weekdays during both experimental weeks. (C) Total sleep time increased during the PRE-INTERVENTION week for good sleepers. Effects between week and weekend among good and poor sleepers are indicated by “**” (*p* ≤ 0.01) and “***” (*p* ≤ 0.001).

### 3.4. Follow-up analysis

One month after the POST-INTERVENTION week, more than 90 % of athletes had pursued to follow at least one sleep recommendation. Interestingly, three months after sleep interventions, poor sleepers maintained more the recommendations than good sleepers (86.4 % vs. 71.4%). For both good and poor sleepers, the protracted sleep intervention effect was found on bedtime (*p* < 0.001, *η*^2^ = 0.37; *p* < 0.001, *η*^2^ = 0.54, respectively). All reported to go to bed earlier one and three months after the sleep interventions compared to PRE-INTERVENTION (*Figure 3*). No difference was found on other variables between PRE-intervention and follow-up data (Table 3 in the supplementary file).

**Figure 3:**
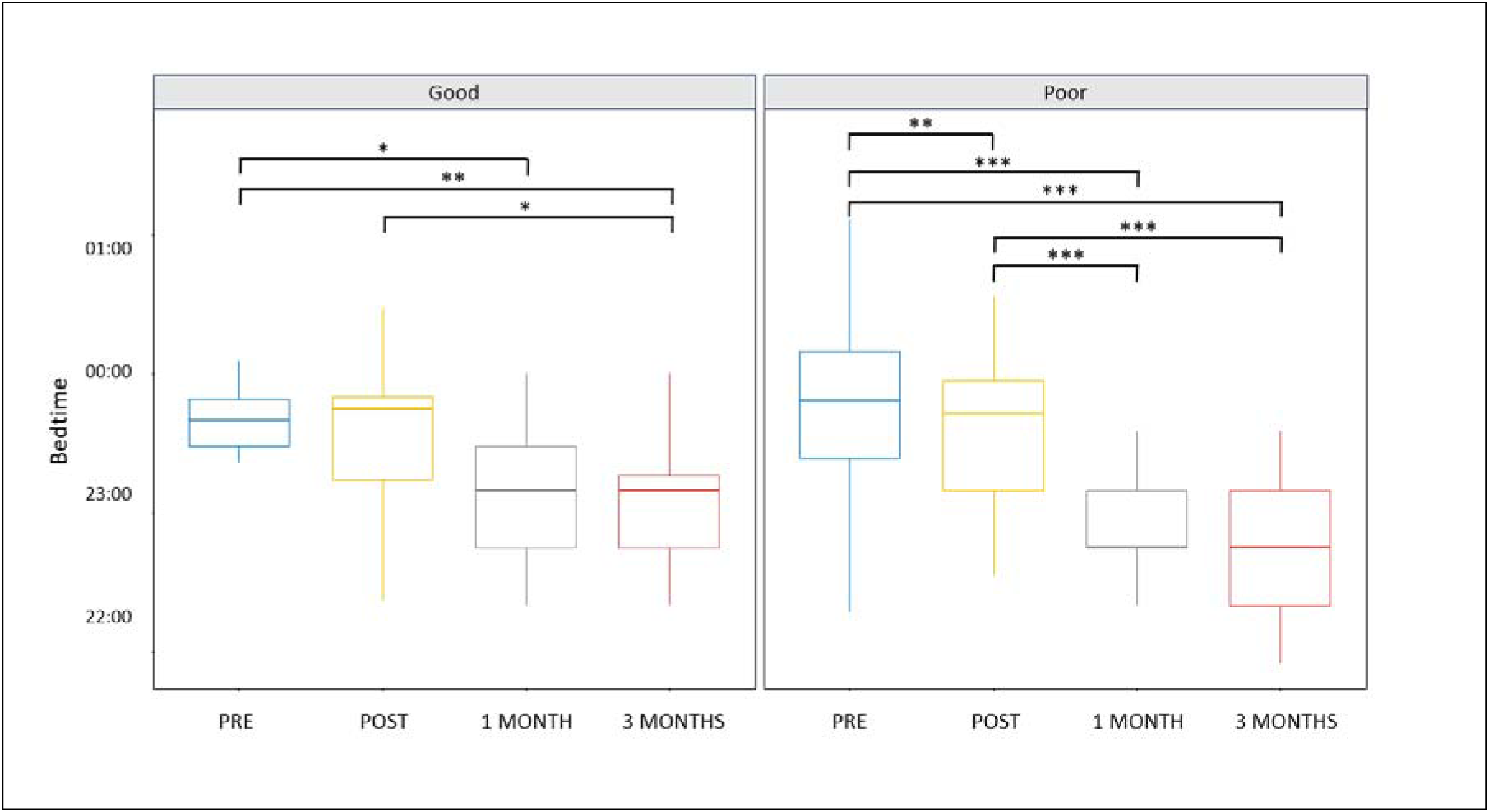
Differences between PRE (blue), POST (yellow), one month post-intervention (grey) and three months post-intervention (red) among good and poor sleepers on bedtime. Effects between week and weekend among good and poor sleepers are indicated by “**” (p ≤ 0.01) and “***” (p ≤ 0.001).

## 4. DISCUSSION

The first aim of this study was to evaluate sleep among team-sport athletes using both objective and subjective assessments identifying good and poor sleepers. The second aim of this study was to examine the contribution of sleep education interventions and relaxation strategies to optimise and enhance sleep in both good and poor sleepers, respectively. Our main results revealed that (i) 61.1 % of rugby players demonstrated poor sleep quality and quantity, (ii) sleep interventions resulted in both objective and subjective sleep improvements, especially among poor sleepers, and (iii) benefits on bedtime remained effective three months after the intervention.

Among the entire group, rugby players exhibited some sleep issues revealing poor sleep quality and quantity, which aligns with previous studies on various athlete populations (Mah et al., 2011; Leeder et al., 2012; Lastella et al., 2015; Swinbourne et al., 2018; Leduc et al., 2019; Vachon et al., 2023). Throughout the PRE-INTERVENTION week, 94.4 % of rugby players slept less than 8 hours per night, including 44 % who slept less than 7 hours per night. Although all rugby players have intermediate or moderate morning chronotype, they went to bed too late (23:43 ± 00:45). This finding had an impact on their amount of TST, which represent a lack of an hour less than the 8 hours recommended for elite athletes (Sargent et al., 2021). Such late bedtime and insufficient sleep quantity appear to be a recurrent observation among rugby players, as previously reported by Lastella et al. (2015) (respectively, 23:43 ± 01:37 and 6.9 ± 1.5 hours). In addition, TST were highly variable between days of the week, hence further demonstrating unregular sleep-wake cycle. This lack of sleep quantity stability observed in our adult athletes was also found and even higher in adolescent rugby players (19.5%; Saidi et al., 2023). Moreover, rugby players went to bed and woke up later during the weekend rather than on weekdays. This discrepancy in the sleep-wake cycle between week and weekend can lead to short-term insufficient sleep quantity that cause acute sleep deprivation and may result in long-term sleep debt (Saidi et al., 2023). This lack of sleep can accumulate over the course of the season and prevent adequate recovery for competition (Lastella et al., 2015). Overall, 61.1 % of rugby players were considered as poor sleepers with the sleep composite score, which aligns with the rate of 50 to 78 % of professional athletes who experience sleep disturbance (Walsh et al., 2021). This multidimensional sleep assessment provides a better understanding of team-sport athletes sleep-wake cycle over a match-week allowing to clearly identify sleep issues to be managed by means of tailored interventions.

The sleep interventions protocol designed here contributed significantly to improve both objective and subjective sleep for poor sleepers. An increase in TST (+ 20.7 min) was found after the interventions, as previously observed after sleep education among rugby players (Caia et al., 2018; Van Ryswyk et al., 2017). This is explained by an earlier bedtime to better match their own chronotype schedule as recommended in the intervention. This earlier bedtime has already been observed among swimmers and resulted in an increase of time in bed (Pasquier et al., 2023). We also found an increase in time in bed (459.7 vs 480.1 min between PRE/POST) but earlier bedtime led here to a longer TST. The lower SE of swimmers in Pasquier et al. (2023) (79.1 % vs 88.8 % in our rugby players) can explain this discrepancy as we may speculate that our relaxation strategies might have led to less sleep fragmentation. Overall, our finding highlights that going to bed earlier, in accordance with one’s own chronotype is an effective recommendation that improves the sleep quantity. During both PRE- and POST-INTERVENTION weeks, wake-up times and bedtimes were delayed on weekends compared to weekdays. This week-end jet lag could lead to an irregular sleep-wake cycle and its removal could be a further approach to pursuit sleep improvement among poor sleepers. Moreover, rugby players demonstrated a daily subjective sleep improvement during the POST-INTERVENTION week, leading to an overall increase in the Hooper score. As this score also includes data on mental well-being, fatigue and muscle soreness, the sleep interventions may have led to a general improvement in well-being, while offering a simple way to recover properly.

Among good sleepers, interventions conducted to better subjective perception of sleep quantity and Hooper score. Such benefit was not previously observed among good sleepers on ESS or monthly subjective sleep assessment (Pasquier et al., 2023). Here, as subjective sleep was daily assessed, its improvement days after days shows that the effects of sleep interventions might be gradually taking hold over time. However, these positive subjective results are compensated by contrasting objective results as has already been described, further reaffirming the need for a multidimensional assessment (Bonnar et al., 2018). Rugby players SE decreased, mainly due to an increase in WASO during the POST-INTERVENTION nights. This objective sleep alteration is associated with an increase of screen time exposure (+ 35 min). As screen time exposure can negatively affect sleep depth (Halson & Juliff, 2017), we suppose that increased exposure to screen time may partially explain the sleep fragmentation of good sleepers during the POST-INTERVENTION week. While bedtime remained stable between the PRE- and POST-INTERVENTION weeks, good sleepers went to bed earlier three months after the interventions (−44 min). Compared to poor sleepers, this change in sleep behaviour for the good sleepers was longer to settle in, which can be explained by the limited gain in their sleep quantity and motivation. As they were considered as good sleepers during the sleep interventions, we suppose that they would have not considered carefully recommendations toward optimisation (i.e., little gain). As sleep is essential for performance and recovery, sleep interventions could be beneficial even for athletes with good sleep assessments.

Even though sleep interventions elicited several positive effects among team-sport players, some limitations should be noted. During the POST-INTERVENTION week, the match took place away from home, and players slept at the hotel on Friday night and took the bus to return home on Saturday evening after the match. Bus discomfort and timing may have delayed bedtime and disrupted their sleep-wake cycle. Moreover, some basic knowledge on sleep is necessary to easily identify athletes’ sleep status using our methodology and intervene specifically to athletes’ needs with simple and basic recommendations. This implies expertise in several sleep dimensions, highlighting the need for sleep education as an integral part of schooling, in the same way as nutrition and physical exercise (Lim et al., 2023).

## 5. PRACTICAL IMPLICATIONS

Coaches, medical staff and sleep experts could use our subjective and objective sleep assessment and intervention program proposed here, by means of questionnaires and actigraphic monitoring. This multidimensional sleep assessment provides a reliable, valid and easily accessible means of monitoring over several weeks, months or years (Claudino et al., 2019). If poor sleep quality is detected, providing sleep interventions could start by explaining simple sleep hygiene recommendations during a sleep education time. A practical part can also be implemented consisting of relaxation techniques such as progressive muscle relaxation, mental imagery and cardiac coherence which was the most practiced one among rugby players. Even more than relaxations techniques, the three recommendations most frequently implemented were napping, going to bed earlier and extending their sleep on the morning. Effects from such sleep interventions can be expected within a week but must be sustained longer to achieve lasting behavioural change necessary to optimise athletes’ health and performance.

## 6. CONCLUSION

The present study confirms that rugby players exhibited poor sleep quality and insufficient sleep quantity (∼7 hours per night) during weekdays and weekends. Sleep interventions including theoretical and practical parts improved players’ objective and subjective sleep characteristics, especially among those with the poorest sleep quality and quantity. Overall, rugby players went to bed earlier, improve their sleep quantity and quality. This multidimensional sleep assessment and interventions proposed here represent a useful and feasible method to improve athletes’ sleep quantity and optimize their sleep quality. As sleep is critical in many physiological and psychological processes, its improvement may be a key factor of performance and injury prevention over a sport season.

## Data Availability

All data produced in the present study are available upon reasonable request to the authors

## ACKWOLEDGMENTS

### Competing interests

The authors declare that they have no competing interests. This project was not supported by any funding source.

### Authors’ contributions

MG contributed to the conceptualization and design the study, interventions on the field and data collection, interpretation of the data, conduction of the statistical analyses, critical drafting and revising the manuscript; AB contributed to the conceptualization and design the study, equipment availability, interpretation of the data and critical revision of the manuscript; BPDF contributed to the conceptualization and design the study, interpretation of the data, and critical revision of the manuscript; JC and VM contributed to data collection and critical revision of the manuscript; ES contributed to the conceptualization and design the study, interventions on the field, interpretation of the data, critical revising the manuscript; UD contributed to the conceptualization and design the study, interpretation of the data, and critical revision of the manuscript. All authors have read and approved the final version of the manuscript and agree with the order of presentation of the authors. Special acknowledgements to Mylan Segaud, master’s student in strengthening and conditioning, who was involved in this project.

## Supplementary Material

### METHOD

#### Sleep interventions procedure

Sleep interventions were separated into two parts. For educational part, there were a group specific feedback among good and poor sleepers. Sleep experts presents their results to each group. For good sleepers, they had to optimize their sleep combining advice on sleep quantity, regularity of bedtime and screen time before bed. For poor sleepers, they had to enhance their sleep focusing on keeping regular sleep-wake cycle, earlier bedtime, increasing sleep quantity (sleep extension in the morning or power nap), adopting a night routine and practicing relaxation strategies, especially before bedtime and during nocturnal awakenings. Sleep experts ended their presentation with the description of the INTERVENTION week tracking.

Thus, practical part began with the explanation of the three relaxation strategies used in mental preparation to help athletes learn to relax and recover mentally and physically. It was explained that these strategies have proven effective in improving sleep, particularly in aiding falling asleep and lengthening deep sleep. They can implement these strategies in combination or individually during INTERVENTION week. The key here was to experience all three techniques, observed their sensitivity to each. We provided them in audio format to reproduce it in self-practice. To begin, they had to lie comfortably on a mat, close their eyes, find the most comfortable position for you, and adjust them. They can already feel the points of contact of their body on the floor… the heels… the calves… the buttocks… the middle of the back… maybe their hands… their elbows… their head… They had to relax as much as possible. The text of strategies is detailed over here :

“Let’s start with the first technique, cardiac coherence… I invite you to focus your attention on your breathing with the goal of matching your breathing rhythm with your heartbeats. Inhale for 5 seconds and exhale deeply for 5 seconds. I invite you to repeat this breathing cycle for 3 minutes while focusing on your breathing… and thinking about nothing in particular… (at 2 minutes) Each cycle enhances the state of relaxation you are in… (at 3 minutes) you can feel more and more harmony between your calm breathing and your internal state calming down, keep going… (At the end of 3 minutes) This is the end of the first exercise.

Now, let’s continue with the second technique, Jacobson’s progressive muscle relaxation… I invite you to follow along with the second technique, which involves alternating cycles of muscle contractions and relaxations in different parts of your body. In each cycle, you will inhale through your nose and gradually contract the indicated muscle area for 5 seconds, then you will perform a brief apnea while still contracting the area, and finally, you will release while exhaling through your mouth. During relaxation, feel the muscle tension release. Breathe deeply, slowly, and at your own pace throughout this exercise.

Let’s start by taking a deep breath, expanding your abdomen. Hold your breath for a few seconds (5-second pause), then exhale slowly through your mouth, feeling like you’re emptying all your air (10-second pause).

Inhale, close your eyes as tightly as possible, contract for 5 seconds… and release the tension as you feel it dissipate (5 seconds)

Next, smile widely, feeling your mouth and cheeks tense up. Hold the tension for about 5 seconds, then release, enjoying the softness of your face (10-second pause)

Now, press your neck against the mat, hold for 5 seconds, then feel the tension as if it were melting away (10-second pause).

Then, feel the weight of your head and neck very light, like a wave flowing from the top of your head to the base of your neck. Inhale and exhale.

Now, focus on your hands, inhale and then without straining, make fists, and maintain this pressure for about 5 seconds, then release (10-second pause).

Now, bend your arms, feel the tension in your biceps. Hold the tension for 5 seconds, then release.

Next, extend your arms by contracting your triceps, feeling your elbows lock in extension. Hold for 5 seconds and release (10-second pause).

Now, contract your trapezius muscles by bringing your shoulders up to your ears.

Hold for 5 seconds and release as the heaviness dissipates (10-second pause)

Try to bring your shoulder blades together, aiming to touch them, hold for 5 seconds, and release (10-second pause).

Then, try to pull in your stomach by contracting your deep abdominal muscles, hold for 5 seconds, and release (10-second pause)

Then, contract your abs and lower back by pressing your lower back against the mat, and feel your lower back against the floor. Hold for 5 seconds and release (10-second pause).

Now, feel a wave of tension starting from the top of your body, your torso, gradually fading away… (5-second pause).

Now, contract your glutes, hold for 5 seconds, then release (10-second pause).

Now squeeze your legs, knee to knee, pressing them together as if holding a pen between your knees (10-second pause).

Then, lift your toes towards your head, keeping your legs straight and heels on the ground, feeling the tension in your calves, hold for 5 seconds, and release as you feel the weight of your legs gradually lift (10-second pause).

Next, point your toes downward, hold for 5 seconds, and release (10-second pause) Now, take a deep breath and feel your lower body relax as you exhale gradually.

Then feel a wave of progressive relaxation starting from the head, passing through the neck, arms, shoulders, and then the trunk, thighs, and finally the feet.

Now, onto the third technique, Mental imagery…

Imagine yourself standing at the crossroads of several paths stretching out before you, each leading to unique destinations… Breathe deeply, and feel the air refreshing your lungs… Become aware of the power of your imagination.

Look at the paths in front of you, each one different, with varied landscapes and promises of a serene and comfortable stroll. Intuitively choose the path that appeals to you the most today, it could be… a green and soothing forest… an idyllic and secluded beach… or a silent mountain offering grand panoramas…

Walk slowly along the path you have chosen, while observing how your body moves in this environment… (proprioceptive)

Explore the details around you… the soft colors… the shapes… the enveloping spaces… (visual)

Listen to the guiding sounds… soothing… close… or more distant… and simply feel the calm and inner peace settling in… (auditory)

As you progress, you may feel a gentle breeze on your skin… smell the scents of the nature around you… (tactile/olfactory)

Each breath is an infusion of tranquility… each exhale is a deeper relaxation…

Engage all your senses and let your body, like your mind, wander freely and peacefully…

Finally, find a comfortable place to sit or lie down in your location…

Feel the softness of this place… Let yourself be rocked by the feeling of relaxation, as if every part of your being were enveloped in soothing warmth…

Allow each breath to deepen your state of relaxation even further…

As you surrender to this inner journey, as you relax even more, let yourself be carried away by regenerative rest…

Take a moment to breathe deeply, fully absorbing the peace of this place… to which you can return at any time when you need it…

Now, as you make your way back…

You can take two or three more dynamic breaths

Open your eyes and return harmonized and invigorated.”

#### Objective and subjective assessments

For objective sleep assessment, the sleep window, defined as the period between the ‘time in bed’ and ‘time out of bed’ was first determined by the algorithm, followed by visual inspection, and verified in concordance with self-reported bed/wake-up times. In the event of a mismatch between these data, a manual correction of the algorithm data was made in favour of the declared bedtime and wake-up times ^21^. For subjective sleep assessment, SDS was a score using five questions on sleep quantity (0 - 4 scale), sleep quality (0 - 4 scale), sleep latency (0 - 3 scale), nocturnal awakenings (0 - 3 scale), and sleep medicine (0 - 3 scale). A total score equal to or below 4 (out of 17) means good sleep. The combination of Hooper sleep score, namely the daily subjective sleep, considers a score of 9 or less out of 19 as indicative of good sleep quality.

### RESULTS

#### Sleep-wake behaviour from initial questionnaire

At the initial questionnaire, analysis of ESS score revealed that seven players had a score between 9 and 14 which means they are sleep-deprived, and three players had a score ≥ 15 which means they had significant general daytime sleepiness. Analysis of MEQ score revealed that 16 players had moderate chronotype (42-58), 17 players are slightly morning type (59-69) and three players are morning type (70-86). Two players reported to take over-the-counter sleep medication at least once a week.

#### PRE- vs POST-INTERVENTION

**Table 1.**
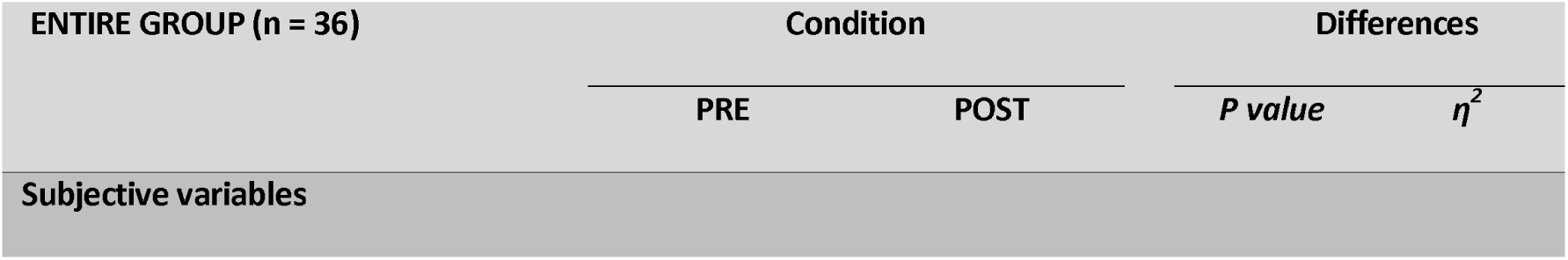
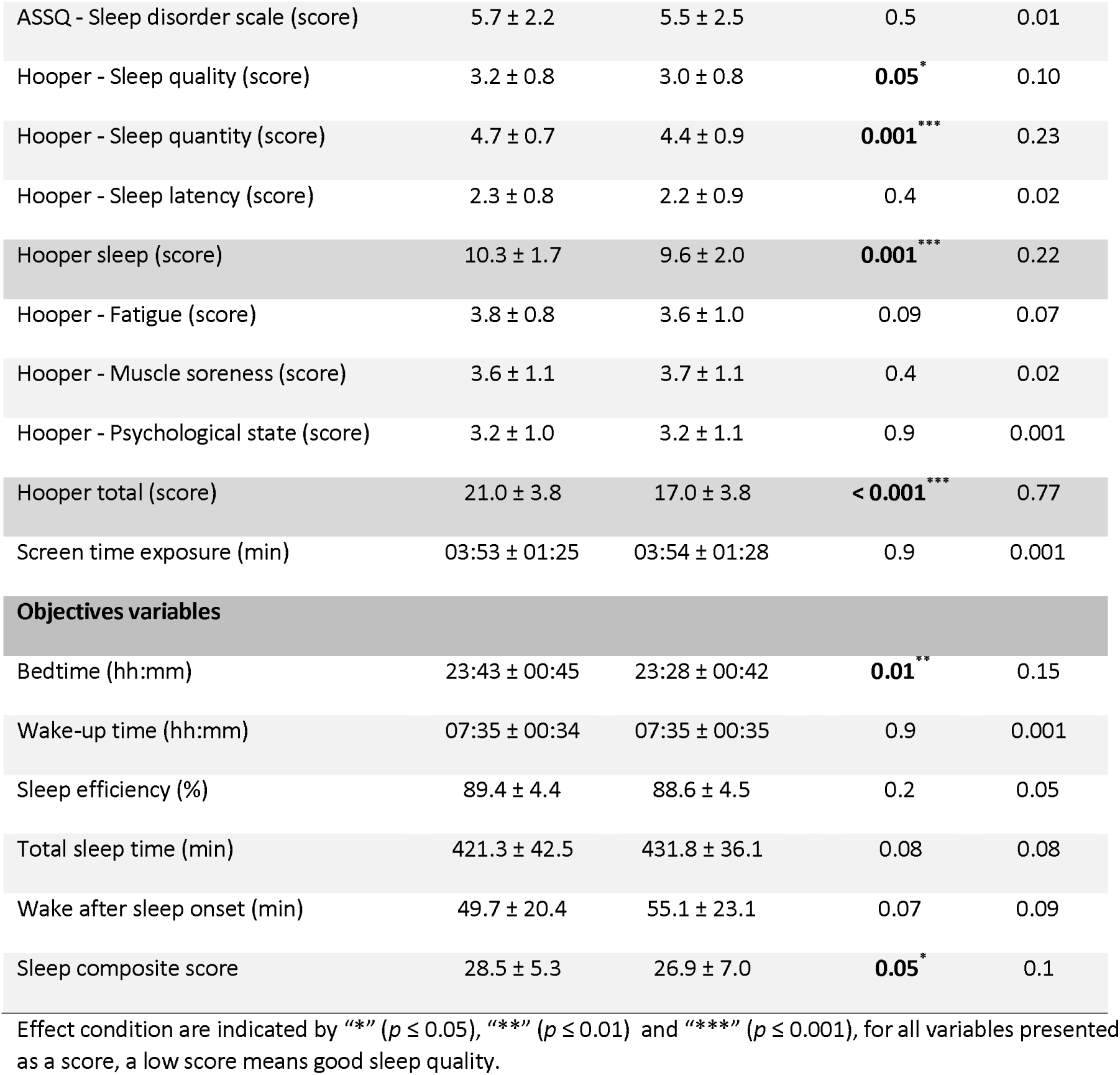
Objective and subjective sleep data throughout the night in each phase for the entire group (OBSERVATION, INTERVENTION; mean and ± SD).

| ENTIRE GROUP (n = 36) | Condition |  | Differences |  |
| --- | --- | --- | --- | --- |
| | PRE | POST | *P value* | $\eta^2$ |
| Subjective variables |  |  |  |  |
| ASSQ - Sleep disorder scale (score) | 5.7 ± 2.2 | 5.5 ± 2.5 | 0.5 | 0.01 |
| Hooper - Sleep quality (score) | 3.2 ± 0.8 | 3.0 ± 0.8 | **0.05\*** | 0.10 |
| Hooper - Sleep quantity (score) | 4.7 ± 0.7 | 4.4 ± 0.9 | **0.001\*\*\*** | 0.23 |
| Hooper - Sleep latency (score) | 2.3 ± 0.8 | 2.2 ± 0.9 | 0.4 | 0.02 |
| Hooper sleep (score) | 10.3 ± 1.7 | 9.6 ± 2.0 | **0.001\*\*\*** | 0.22 |
| Hooper - Fatigue (score) | 3.8 ± 0.8 | 3.6 ± 1.0 | 0.09 | 0.07 |
| Hooper - Muscle soreness (score) | 3.6 ± 1.1 | 3.7 ± 1.1 | 0.4 | 0.02 |
| Hooper - Psychological state (score) | 3.2 ± 1.0 | 3.2 ± 1.1 | 0.9 | 0.001 |
| Hooper total (score) | 21.0 ± 3.8 | 17.0 ± 3.8 | **< 0.001\*\*\*** | 0.77 |
| Screen time exposure (min) | 03:53 ± 01:25 | 03:54 ± 01:28 | 0.9 | 0.001 |
| **Objectives variables** |  |  |  |  |
| Bedtime (hh:mm) | 23:43 ± 00:45 | 23:28 ± 00:42 | **0.01\*\*** | 0.15 |
| Wake-up time (hh:mm) | 07:35 ± 00:34 | 07:35 ± 00:35 | 0.9 | 0.001 |
| Sleep efficiency (%) | 89.4 ± 4.4 | 88.6 ± 4.5 | 0.2 | 0.05 |
| Total sleep time (min) | 421.3 ± 42.5 | 431.8 ± 36.1 | 0.08 | 0.08 |
| Wake after sleep onset (min) | 49.7 ± 20.4 | 55.1 ± 23.1 | 0.07 | 0.09 |
| Sleep composite score | 28.5 ± 5.3 | 26.9 ± 7.0 | **0.05\*** | 0.1 |

1 Effect condition are indicated by “\*” ( $p \leq 0.05$ ), “\*\*” ( $p \leq 0.01$ ) and “\*\*\*” ( $p \leq 0.001$ ), for all variables presented 2 as a score, a low score means good sleep quality.

#### Week days and weekend analysis

During the PRE-INTERVENTION week, good sleepers woke up later during the weekend rather than during the week days, inducing greater TST (*p* ≤ 0.001, *η*^2^ = 0.11 and *p* = 0.02, *η*^2^ = 0.04 respectively) (*Figure 1 in supplementary* ). No difference was found between week and weekend nights among good sleepers during the POST-INTERVENTION week (Table 2 in supplementary file). During PRE-INTERVENTION week, poor sleepers went to bed and woke up later during weekend rather than weekdays (*p* ≤ 0.001, *η*^2^ = 0.07 and *p* ≤ 0.001, *η*^2^ = 0.15 respectively) inducing no increase in TST (*p* = 0.4) (Table 2 in supplementary file). They conserved the same shift observed in PRE-INTERVENTION week going to bed and waking up later during the weekend rather than weekdays (*p* = 0.007, *η*^2^ = 0.07; *p* = 0.003, *η*^2^ = 0.08 respectively) while TST remained unchanged (*p* = 0.8) during POST-INTERVENTION week.

**Figure 1:**
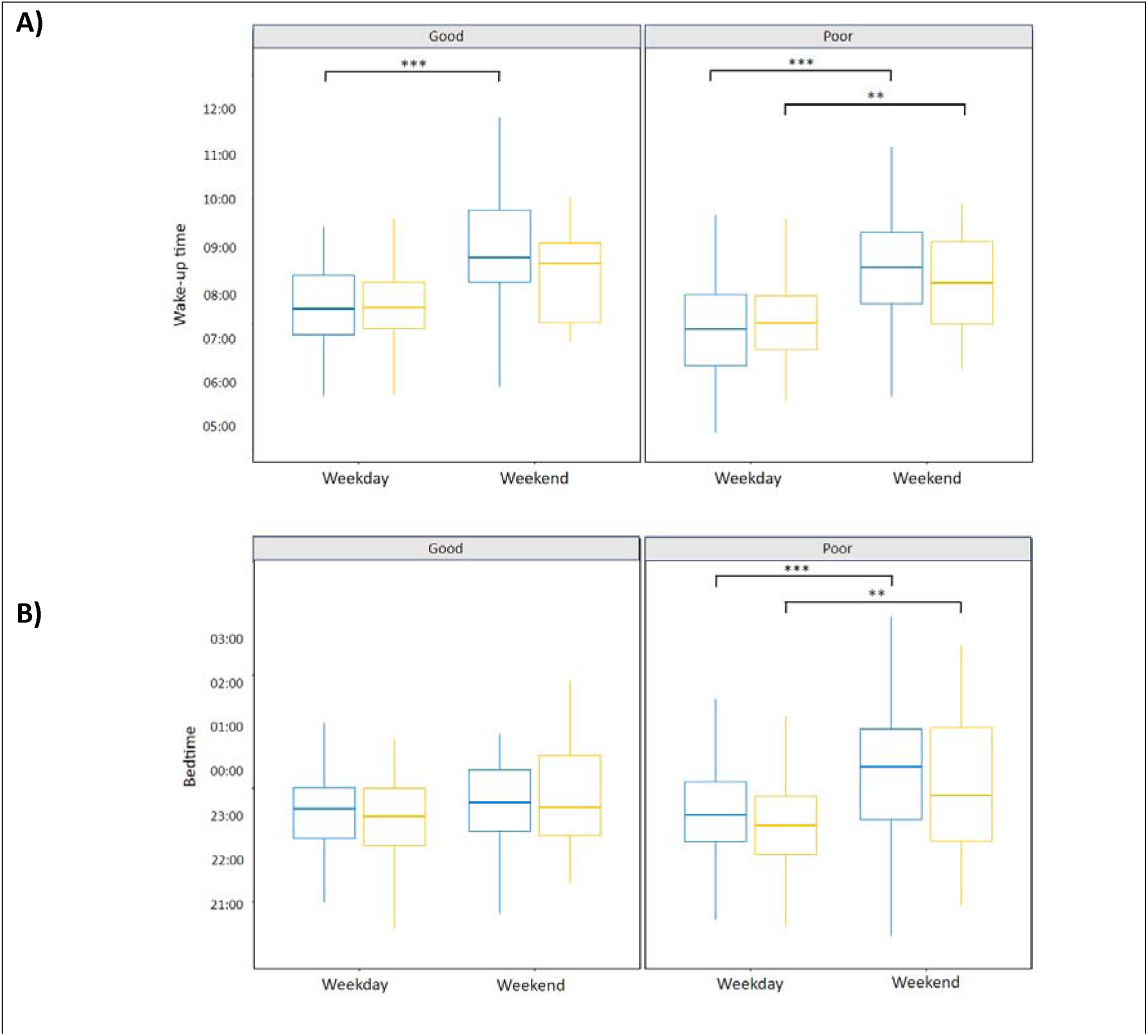
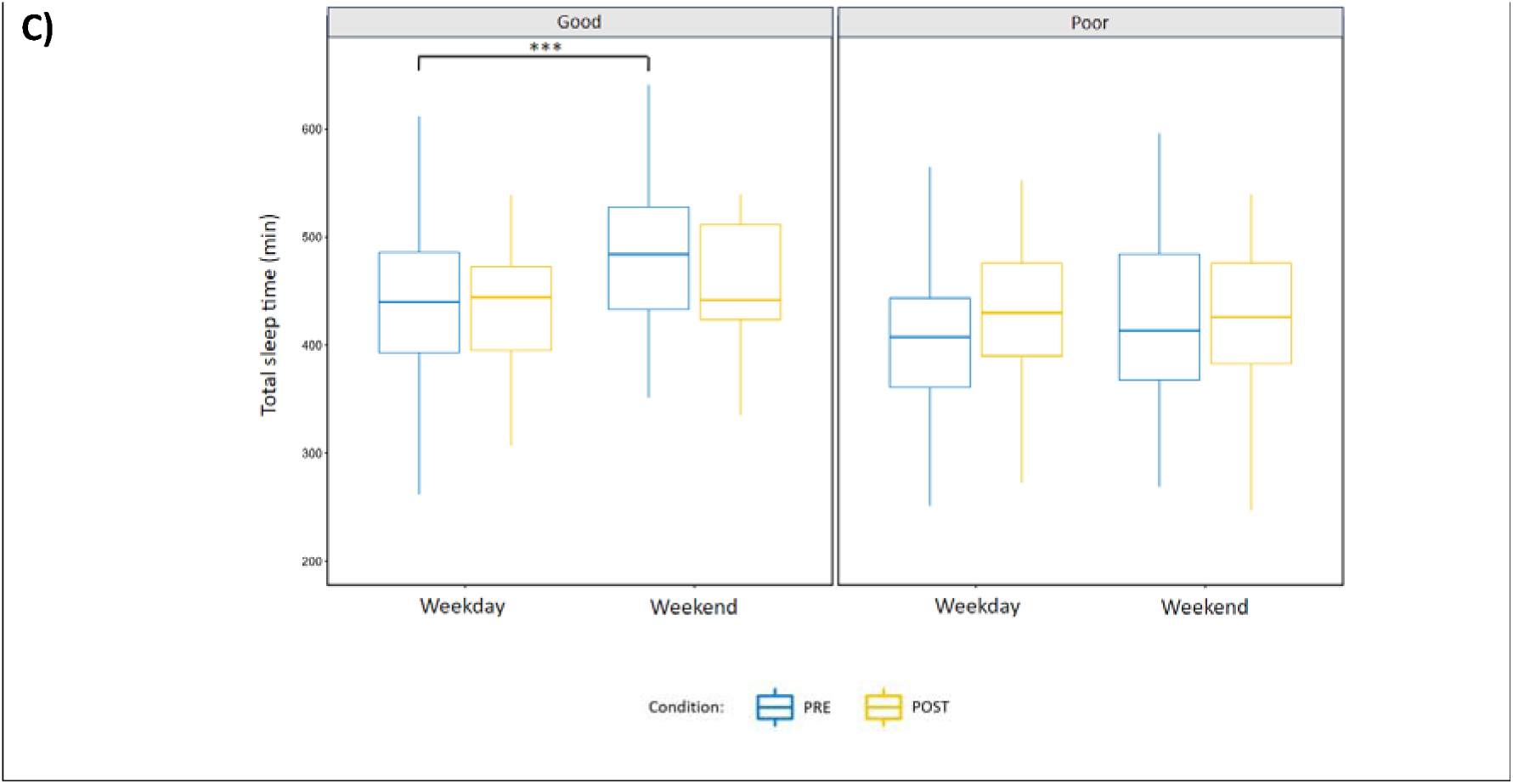
Differences between week and weekend nights (Friday / Saturday) among good and poor sleepers on wake-up time (A), bedtime (B), total sleep time (C) during PRE- (blue) and POST-INTERVENTION (yellow) weeks. (A) Good and poor sleepers woke up later during the weekend compared to weekdays during PRE- and POST-INTERVENTION only for poor sleepers. (B) Poor sleepers went to bed later during the weekend compared to weekdays during both experimental weeks. (C) Total sleep time increased during the PRE-INTERVENTION week for good sleepers. Effects between week and weekend among good and poor sleepers are indicated by “**” (p ≤ 0.01) and “***” (p ≤ 0.001).

**Table 2.** Evaluation of difference between week and weekend nights during PRE-INTERVENTION among the entire group, good sleepers and poor sleepers (mean ± SD).

| PRE-INTERV. | ENTIRE GROUPE (n=36) |  |  | GOOD SLEEPERS (n = 14) |  |  | POOR SLEEPERS (n = 22) |  |  |
| --- | --- | --- | --- | --- | --- | --- | --- | --- | --- |
|  | Week | Weekend | *P value* | Week | Weekend | *P value* | Week | Weekend | *P value* |
| BT (hh:mm) | 23:32 ± 01:11 | 00:13 ± 01:35 | .001*** | 23:32 ± 00:57 | 23:40 ± 00:54 | .4 | 23:32 ± 01:19 | 00:31 ± 01:47 | <.001*** |
| WUT (hh:mm) | 07:18 ± 00:54 | 08:26 ± 01:25 | <.001*** | 07:31 ± 00:52 | 08:35 ± 00:52 | <.001*** | 07:09 ± 00:55 | 08:21 ± 01:29 | <.001*** |
| SE (%) | 89.5 ± 6.5 | 89.8 ± 7.0 | .7 | 90.9 ± 4.9 | 89.2 ± 7.3 | .3 | 88.6 ± 7.2 | 90.1 ± 6.8 | .2 |
| TST (min) | 416.7 ± 71.6 | 442.2 ± 87.2 | .05 | 435.7 ± 67.8 | 479.3 ± 74.9 | .02* | 403.8 ± 71.6 | 421.8 ± 87.6 | .4 |
| WASO (min) | 49.0 ± 32.1 | 50.1 ± 35.9 | .9 | 43.0 ± 23.1 | 56.0 ± 36.7 | .1 | 53.0 ± 36.5 | 46.9 ± 35.5 | .3 |
| Hopper sleep | 10.4 ± 2.6 | 10.1 ± 2.7 | .3 | 9.0 ± 2.3 | 8.3 ± 2.2 | .08 | 11.3 ± 2.3 | 11.2 ± 2.4 | .9 |
| Screen | 03:55 ± 01:47 | 03:36 ± 01:37 | .2 | 03:55 ± 01:38 | 03:26 ± 01:30 | .2 | 03:54 ± 01:54 | 03:42 ± 01:42 | .6 |

Effect between Week and Weekend among good and poor sleepers are indicated by “\*” ( $p \leq 0.05$ ) and “\*\*\*” ( $p \leq 0.001$ ).

**Table 3.**
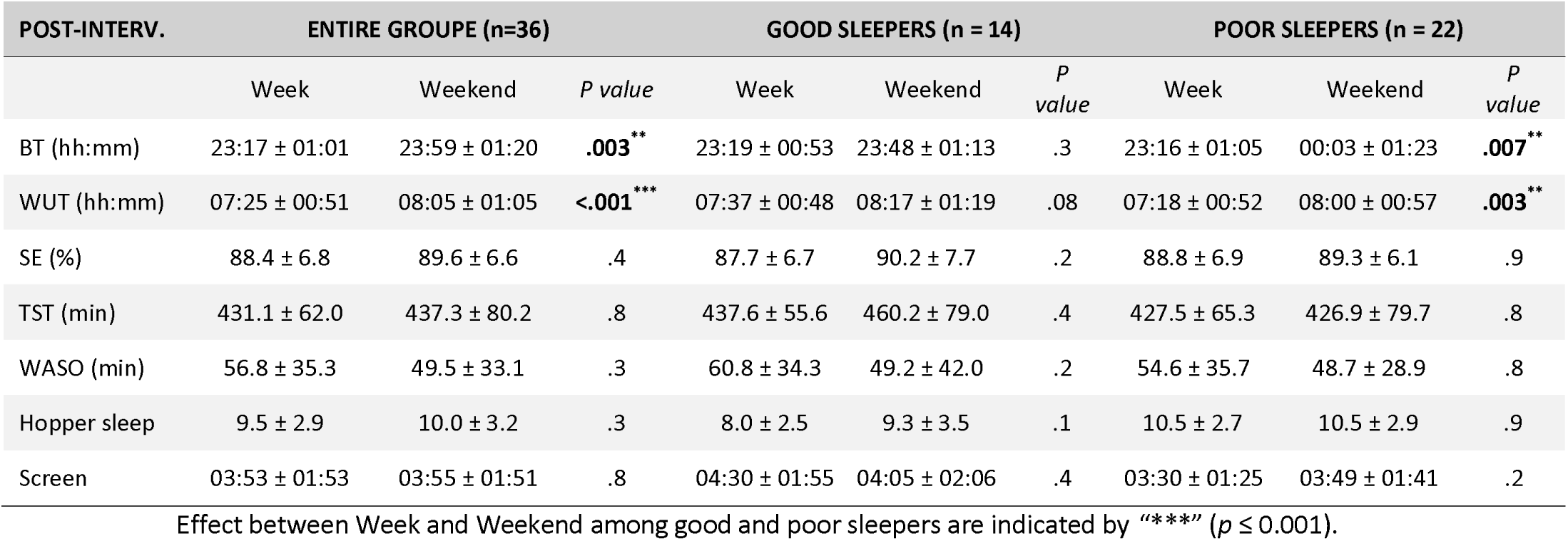
Evaluation of difference between week and weekend nights during INTERVENTION among the entire group, good sleepers and poor sleepers (mean ± SD).

| POST-INTERV. | ENTIRE GROUPE (n=36) |  |  | GOOD SLEEPERS (n = 14) |  |  | POOR SLEEPERS (n = 22) |  |  |
| --- | --- | --- | --- | --- | --- | --- | --- | --- | --- |
|  | Week | Weekend | *P value* | Week | Weekend | *P value* | Week | Weekend | *P value* |
| BT (hh:mm) | 23:17 ± 01:01 | 23:59 ± 01:20 | **.003\*\*** | 23:19 ± 00:53 | 23:48 ± 01:13 | .3 | 23:16 ± 01:05 | 00:03 ± 01:23 | **.007\*\*** |
| WUT (hh:mm) | 07:25 ± 00:51 | 08:05 ± 01:05 | **<.001\*\*\*** | 07:37 ± 00:48 | 08:17 ± 01:19 | .08 | 07:18 ± 00:52 | 08:00 ± 00:57 | **.003\*\*** |
| SE (%) | 88.4 ± 6.8 | 89.6 ± 6.6 | .4 | 87.7 ± 6.7 | 90.2 ± 7.7 | .2 | 88.8 ± 6.9 | 89.3 ± 6.1 | .9 |
| TST (min) | 431.1 ± 62.0 | 437.3 ± 80.2 | .8 | 437.6 ± 55.6 | 460.2 ± 79.0 | .4 | 427.5 ± 65.3 | 426.9 ± 79.7 | .8 |
| WASO (min) | 56.8 ± 35.3 | 49.5 ± 33.1 | .3 | 60.8 ± 34.3 | 49.2 ± 42.0 | .2 | 54.6 ± 35.7 | 48.7 ± 28.9 | .8 |
| Hopper sleep | 9.5 ± 2.9 | 10.0 ± 3.2 | .3 | 8.0 ± 2.5 | 9.3 ± 3.5 | .1 | 10.5 ± 2.7 | 10.5 ± 2.9 | .9 |
| Screen | 03:53 ± 01:53 | 03:55 ± 01:51 | .8 | 04:30 ± 01:55 | 04:05 ± 02:06 | .4 | 03:30 ± 01:25 | 03:49 ± 01:41 | .2 |

Effect between Week and Weekend among good and poor sleepers are indicated by “\*\*\*” ( $p \leq 0.001$ ).

#### Evaluation of sleep interventions

**Table 4.**
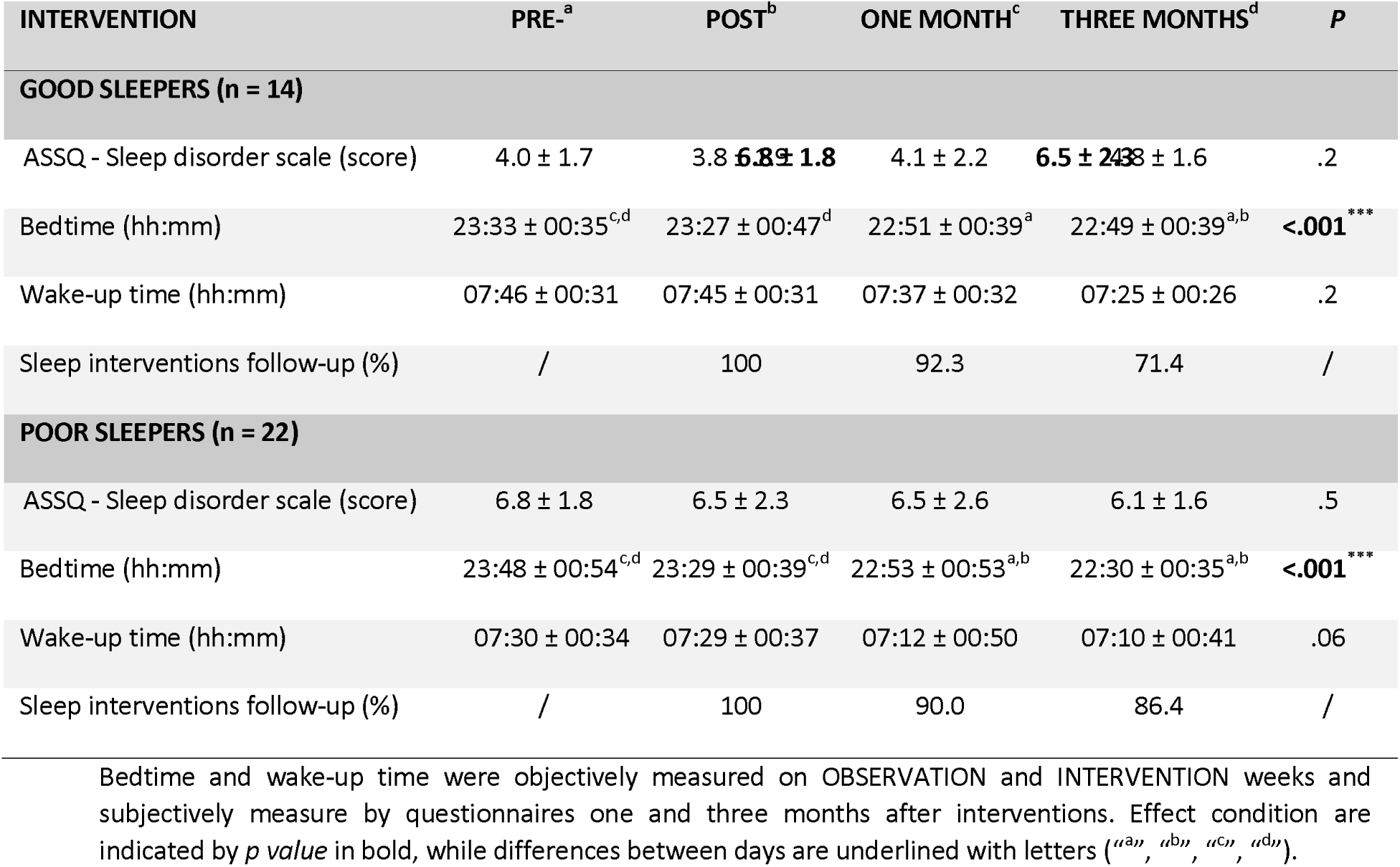
Evaluation of sleep intervention over three months.

## Notes

### Competing Interest Statement

The authors have declared no competing interest.

### Funding Statement

This study did not receive any funding

### Author Declarations

Ethical committee of Hospices Civil Lyon gave approval for this work (23-112).

